# Solid-Phase Extraction and Enhanced Amplification-Free Detection of Pathogens Integrated by Dual-Functional CRISPR-Cas12a

**DOI:** 10.1101/2023.04.28.23289279

**Authors:** Zimu Tian, He Yan, Yong Zeng

## Abstract

Public healthcare demands effective and pragmatic diagnostic tools to address the escalating challenges in infection management in resource-limited areas. Recent advance in CRISPR-based biosensing promises the development of next-generation tools for disease diagnostics, including point-of-care (POC) testing for infectious diseases. Currently prevailing strategy of developing CRISPR assays exploits only the non-specific trans-cleavage function of a CRISPR-Cas12a/Cas13a system for detection and combines it with an additional pre-amplification reaction to enhance the sensitivity. In contrast to this single-function strategy, here we present a new approach that collaboratively integrates the dual functions of CRISPR-Cas12a: sequence-specific binding and trans-cleavage activity. With this approach, we developed a POC nucleic acid assay termed Solid-Phase Extraction and Enhanced Detection assay Integrated by CRISPR-Cas12a (SPEEDi-CRISPR) that negates the need for preamplification but significantly improves the detection of limit (LOD) from the pM to fM level. Specifically, using Cas12a-coated magnetic beads, this assay combines efficient solid-phase extraction and enrichment of DNA targets enabled by the sequence-specific affinity of CRISPR-Cas12a with the fluorogenic detection by the activated Cas12a on beads. Our proof-of-concept study demonstrated that the SPEEDi-CRISPR assay affords an improved detection sensitivity for human papillomavirus (HPV)-18 with a LOD of 2.3 fM and excellent specificity to discriminate HPV-18 from HPV-16, Parvovirus B19, and scramble HPV-18. Furthermore, this robust assay was readily coupled with a portable smartphone-based fluorescence detector and a lateral flow assay for quantitative detection and visualized readout, respectively. Overall, these results should suggest that our dual-function strategy could pave a new way for developing the next-generation CRISPR diagnostics and that the SPEEDi-CRISPR assay provides a potentially useful tool for point-of-care testing.

## Introduction

Infectious diseases pose a major threat to public health, such as the recent COVID-19 pandemic associated with SARS-CoV-2 and human papillomavirus (HPV) that may lead to cervical cancers. Techniques for sensitive and rapid detection of pathogens are essential to prevent complications arising out of the disease progression and are beneficial for formulating an effective therapy for patients.^1-6^ Nucleic acid testing technologies led by PCR pave the way for the modern pathogen identification owning to its powerful ability of signal amplification. More recently, the emerging CRISPR (clustered regularly interspaced short palindromic repeats)-Cas (CRISPR-associated) systems such as CRISPR-Cas12/13 have been demonstrated as the next-generation biosensing technology^7-12^ and have drawn increasing interest in the development of isothermal diagnostic assays.^13-15^ CRISPR-based detection involves the specific binding of targets to the Cas enzyme-CRISPR RNA (crRNA) ribonucleoprotein (RNP) complex and the subsequent signal amplification via the trans-cleavage of fluorogenic reporters.^7, 16-17^ Though CRISPR-Cas has been applied to the development of various detection systems, ^18-22^ strategies that only rely on the trans-cleavage activity suffer from limited sensitivity in clinical detection due to the low concentration of targets, thus, current CRISPR diagnostics (CRISPR-Dx) requires additional nucleic acid amplification techniques, such as loop-mediated isothermal amplification (LAMP)^23-24^ and recombinase polymerase amplification (RPA)^25-26^ to generate sufficient amplicons for RNP. However, the pre-amplification raises the reagent complexity and the risk of contamination during multiple manual steps. So, an amplification-free CRISPR-Cas assay with high sensitivity would greatly benefit CRISPR-Dx.

It is worth noting that even if the volume of biological samples consumed for laboratory testing has been dramatically reduced now, the volume of biological samples collected for diagnosis, such as blood,^27-29^ saliva,^30^ and swab media,^31^ typically exceeds the volume needed when taking sample storing and transporting into account. In most cases, a majority of target molecules in collected samples are wasted, which restricts the potential analytical performance that a bioassay might achieve. Hence, the extraction and enrichment of target molecules from abundant biological samples would greatly benefit amplification-free detection.

Herein, we report a Solid-Phase Extraction and Enhanced Detection assay Integrated by CRISPR-Cas12a (SPEEDi-CRISPR) for amplification-free and sensitive analysis of pathogen nucleic acids from biological samples. The Cas12a-crRNA RNP is conjugated on the surface of magnetic beads via the affinity between the poly-histidine tags on Cas12a and the cobalt ions on the beads. As illustrated in **Fig. 1A**, target DNAs in samples will first be captured on beads via the sequence recognition of Cas12a-crRNA RNP, which will thereafter activate the trans-cleavage function of Cas12a (**Fig. 1B**). Then, after removal of the supernatant and addition of the detection buffer, signals will be generated by the cleavage of ssDNA reporters. The fluorescence signals of SPEEDi-CRISPR can be quantified in a bench-top instrument such as the qPCR apparatus or plate reader, alternatively, it could be coupled with a smartphone-based portable device or lateral flow assay (LFA) for a visualized readout to meet the demand for point-of-care testing (POCT). Our SPEEDi-CRISPR offers three major distinctions from the existing CRISPR-Dx. First, it leverages the affinity between the Cas12a-crRNA RNP and its DNA target, which is used as a solid-phase extraction strategy for the first time. Second, our amplification-free assay affords superior analytical performance in detecting pathogen DNA with high sensitivity (single-digit fM) and excellent specificity. Third, our method integrates target-specific capture, sample enrichment, and signal generation into one single material, drastically simplifying the assay workflow, decreasing the risks of false signals caused by contamination, and reducing the reliance on equipment and reagents.

**Figure 1.**
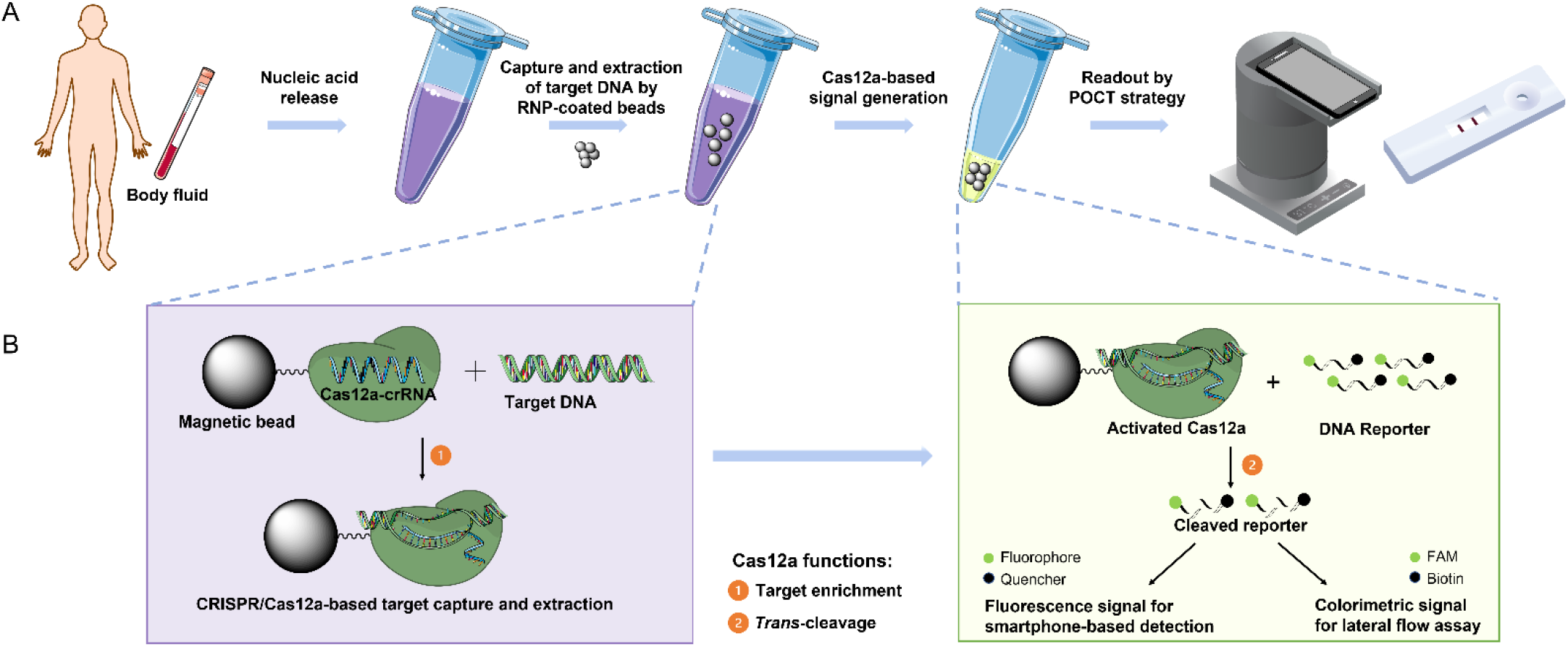
SPEEDi-CRISPR assay. **(A)** Workflow of SPEEDi-CRISPR for POC diagnosis. Nucleic acids are released via heat-lysis of samples; then Cas12a-crRNA RNP-coated beads are added to a large volume of samples for the specific capture and enrichment of target DNA. After enrichment, the supernatant is discarded and the reaction buffer containing reporters is added for signal generation. The result can be measured in a bench-top instrument, it can also be coupled with a portable device or LFA for visualized readout in POCT. **(B)** Principle of SPEEDi-CRISPR assay. Cas12a-crRNA RNP is coated on the beads via the coordination between the His-tag on Cas12a’s N-terminal and the cobalt ions on the magnetic beads. Functionalized beads capture target DNA by sequence recognition of Cas12a-crRNA RNP. After extraction, the activated Cas12a will cleave reporters in the reaction buffer for signal generation.

As a proof-of-concept for potential applications in diagnosis, we investigated the ability of SPEEDi-CRISPR to detect the two high-risk HPV subtypes (HPV 16 and HPV 18) which account for roughly 70% of all cervical cancers.^32-33^ In addition, lateral flow assay and a homemade portable device have been coupled with our assay and can successfully detect the two high-risk HPV subtypes. We believe that our SPEEDi-CRISPR assay is promising to advance the POCT of cervical cancers with minimal use of complex reagents and instruments, particularly in resource-limited areas and developing countries.

## Results and Discussion

### Feasibility test of the SPEEDi-CRISPR assay

The interaction between the Cas12a-crRNA RNP and its target DNA has great specificity and strong binding affinity (dissociation constant *K*_d_=3.9 nM for LbCas12a).^34-37^ We believe this target binding affinity can be leveraged to design a solid-phase extraction material for the specific enrichment of DNA targets. The fact that the trans-cleavage of Cas12a is activated after target binding makes it possible for the subsequent signal generation,^37^ hyphenating the extraction and detection in a single functionalized material. We first confirmed the conjugation of RNPs on the surface of magnetic beads. The Cas12a-crRNA complex was prepared using the procedure mentioned in methods and coated on dynabeads with the maximum coating capacity (1.4 pmol Cas12a on 1 μg beads). As shown in **Fig. S1**, the Cas12a-crRNA RNP modification decreased the zeta potential of beads in 1× PBS buffer from -40 mV to -60 mV, showing that the Cas12a-crRNA complex was modified successfully on the bead surface via the interaction between the his-tag and cobalt ions.

Next, using the L1 locus^7^ (**Fig. S2**) in the HPV genome as the model target, the activity of the Cas12a-crRNA complex on the beads was verified. In our design, the trans-cleavage activity of RNPs on beads will be initiated once the RNP captures the target DNA in solutions, and the cleavage of the fluorogenic ssDNA reporter will generate fluorescence signals. In the experiment, the RNPs or RNP-coated beads were added into the reaction buffer containing 50 nM reporter. In the presence of target DNA, an increase in the fluorescence intensity was observed (**Fig. 2A**), and relatively weak background fluorescence signals were monitored in the absence of the target DNA, confirming that the target DNA triggers the activation of the trans-cleavage function of Cas12a. RNP-modified beads also displayed a signal increase with target DNA added while the unmodified beads in the solution containing target DNA did not clearly show signal enhancement compared to the background, demonstrating that the RNPs on beads contribute to the fluorescence signals (**Fig. 2A**). The experiment verified that the sequence recognition and trans-cleavage activities of RNP are retained after conjugation and could be utilized for target capture and the following signal generation. However, the fluorescence intensity from RNPs coated on beads is slightly lower than that from the RNPs in free solution, indicating the limited activity of RNP on beads in this reaction condition which needs further optimizations. We furthermore tested the extraction ability of the RNP-coated beads. The beads were used to extract the 100-μL sample containing 10 nM targets, and the end-point fluorescence signal from the cleaved reporters was measured after extraction. As shown in **Fig. 2B**, the signal using our beads is greater than that of the non-extraction experiment where only 2 μL of the sample was used, suggesting the successful enrichment of target DNA in the SPEEDi-CRISPR assay. Overall, the SPEEDi-CRISPR assay is able to enrich targets from large-volume samples to produce higher fluorescence signals, indicating that our strategy has the potential to be developed into a preamplification-free nucleic acid detection method.

**Figure. 2.**
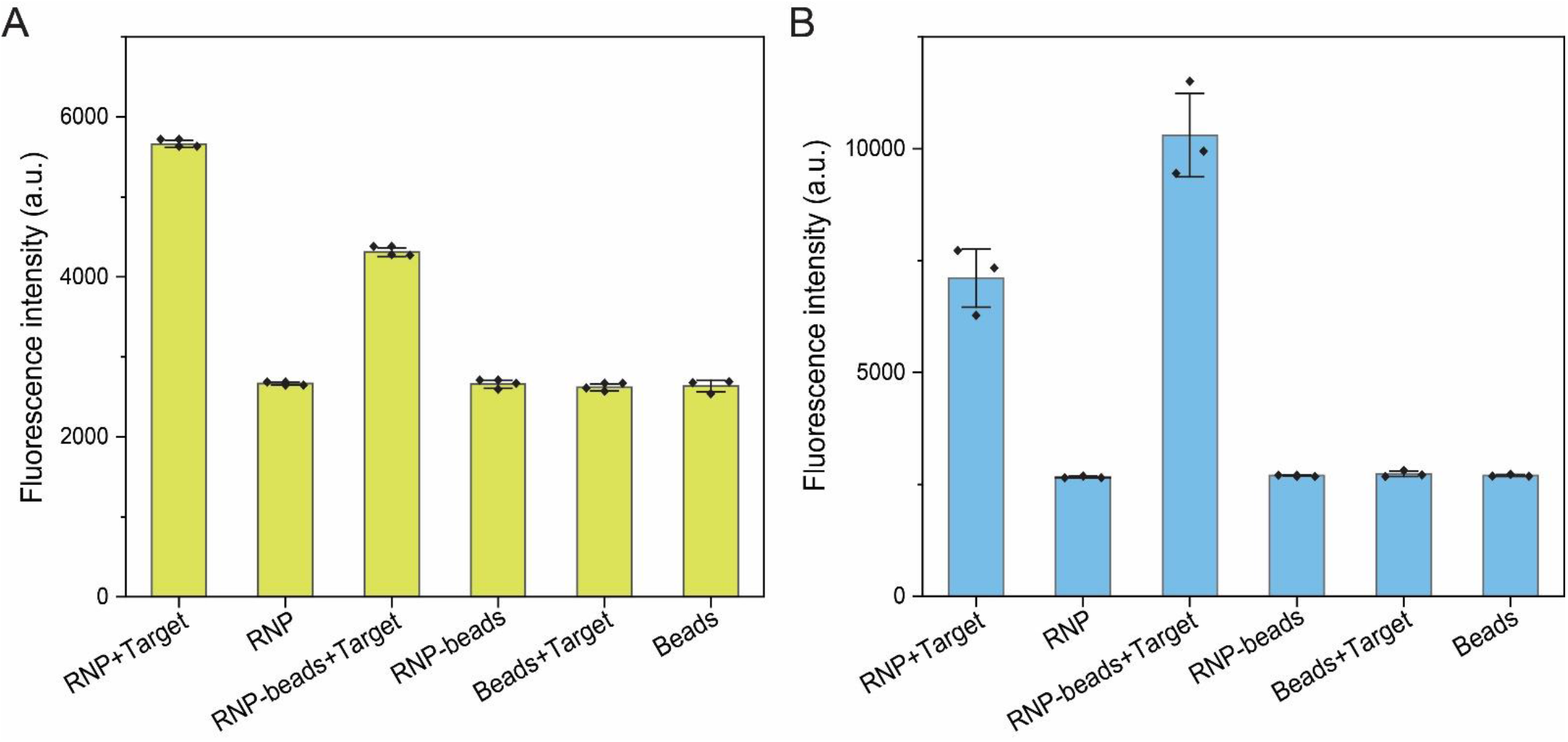
Feasibility test of SPEEDi-CRISPR assay. **(A)** Validation of trans-cleavage activity of Cas12a on beads. Only the RNP + Target group and the RNP-Beads + Target group showed increased fluorescence intensity compared to the background signal, suggesting that Cas12a modified on beads retains the trans-cleavage activity. All measurements were performed by a qPCR device. Error bars: one S.D. (*n* = 3). **(B)** Target extraction using functionalized beads. After extraction, more targets were captured on the beads, resulting in a higher signal (RNP-beads + Target) compared to the non-extraction experiment (RNP + Target). All measurements in this figure were performed by a qPCR device. Error bars: one S.D. *(n* = 3).

### Optimization of the SPEEDi-CRISPR assay

We systematically investigated several major factors that affect the performance of the SPEEDi-CRISPR assay. The amount of RNP modified on the bead surface is a key variable in our assay as it will impact both target enrichment and subsequent signal generation in the solid-phase reaction format. As shown in **Fig. 3A**, SPEEDi-CRISPR produced the highest fluorescence in the detection of 1 pM target when 1.25 pmol of RNPs were coated on per μg of beads. The amount of RNPs-coated beads used in the assay has been investigated as well (**Fig. S3A**), however, we didn’t see any significant changes in signal intensity when the input bead quantity increased from 20 μg to 60 μg, so we selected 20 μg for the following optimization. Next, the concentration of the reporter in the reaction buffer was tested. We screened a wide range of reporter concentrations from 100 to 800 nM and the optimal concentration of the reporter was chosen at 600 nM (**Fig. 3B**) since it displayed the highest signal. Furthermore, operations in the extraction and signal generation step of the SPEEDi-CRISPR assay have been optimized. Based on the outcomes of these tests, the ideal extraction time (25 min, **Fig. 3C**) and reaction time (80 min, **Fig. 3D**) for the SPEEDi-CRISPR were established, allowing the assay to be completed in 2 hours. Moreover, the SPEEDi-CRISPR assay can benefit from multiple rounds of extraction to improve sensitivity further when the volume of samples is abundant. As shown in **Fig. S3B**, fluorescence signals may be raised by increasing the number of extraction rounds, but they started to decline after the fourth round, which may be related to the reduced trans-cleavage capacity of Cas12a because of the prolonged extraction process. In further tests, only one round of extraction would be utilized for a fast turnaround time. The aforementioned optimization led to the establishment of a standard SPEEDi-CRISPR assay protocol.

**Figure 3.**
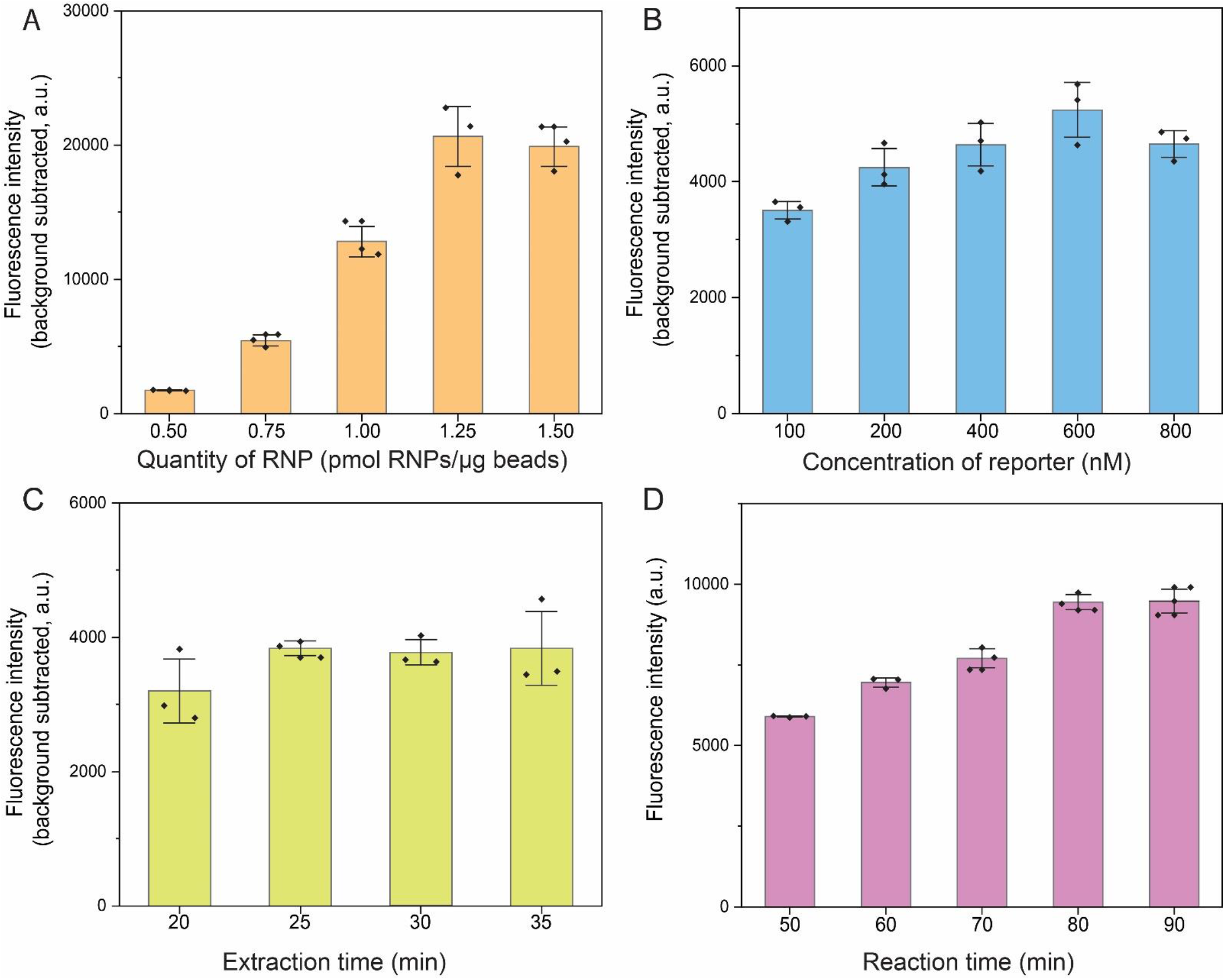
Optimization of SPEEDi-CRISPR. **(A)** Optimization of the RNP amount on the surface of magnetic beads. Increasing the amount of RNP from 0.5 to 1.25 pmol/μg beads could increase the fluorescence intensity, suggesting that 1.25 pmol/μg beads is the optimal condition. **(B)** Optimization of reporter concentrations. The fluorescence intensity increases when the reporter concentration is increased from 100 nM to 600 nM. Inhibition occurs when the concentration is more than 600 nM, suggesting that 600 nM is the optimal concentration. **(C)** Optimization of extraction time. The fluorescence intensity increases when the extraction time rises from 20 to 25 minutes but does not change when the extraction time is extended beyond 25 minutes, suggesting that 25 minutes is the ideal extraction time. **(D)** Optimization of reaction time for signal generation. When the reaction time rises from 20 min to 100 min, the fluorescence intensity increases from 20 min to 80 min and reaches the plateau when the reaction time exceeds 100 min, implying that 80 min is the best reaction time. All measurements were performed by a qPCR device. Error bars: one S.D. (*n* = 3, 4, or 5).

### Characterization of the SPEEDi-CRISPR assay

As discussed above, SPEEDi-CRISPR is expected to achieve the enhanced amplification-free detection of target DNA via target enrichment. We first investigated the limit of detection (LOD) of the SPEEDi-CRISPR assay for the quantitative detection of HPV-18 DNA under optimized experimental conditions. The beads were used to extract target DNAs from the samples (100 μL) with varying concentrations and fluorescence signals were evaluated (**Fig. 4A**). There was an excellent linear relationship between the fluorescence signal and the concentration of target DNA in the range of 1 pM to 5 fM (R^2^ = 0.995) (**Fig. 4B**). The LOD for HPV-18 testing was determined to be 2.3 fM based on the 3σ rule. Meanwhile, these samples were also tested using the CRISPR-Dx method without extraction (**Fig. S4A and C**). The results showed that conventional CRISPR-Dx without extraction can only achieve detection sensitivity of HPV-18 DNA at pM-level (**Fig. S4A**), demonstrating that the SPEEDi-CRISPR assay can significantly enhance the sensitivity of CRISPR-Dx. We next evaluated the specificity of our assay. CrRNA in the assay specifically designed for HPV-18 was used to identify samples containing 10 nM HPV-18, HPV-16, Parvovirus B19 (PB 19), and the scrambled sequence of HPV-18 target DNA. As shown in **Fig. 4C**, the non-targets yielded a signal level that has no significant difference from the negative control, whereas samples containing HPV-18 DNA generated a considerably high signal, demonstrating excellent specificity of the SPEEDi-CRISPR assay to discriminate HPV-18 from other non-targets.

**Figure 4.**
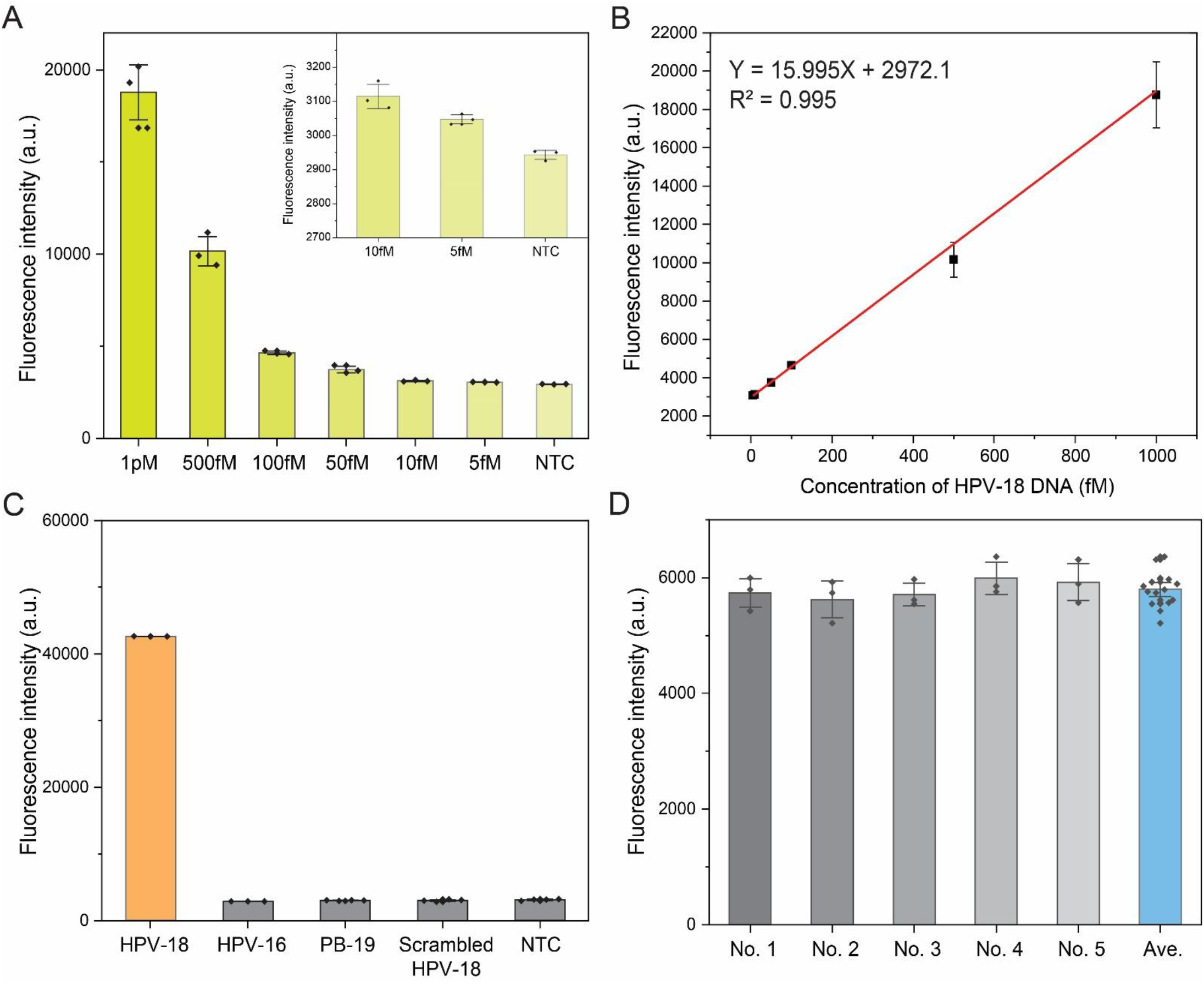
Analytical performance of SPEEDi-CRISPR in the detection of HPV-18. **(A)** Fluorescence intensity of experiments with different target concentrations in optimal conditions. Error bars: one S.D. (*n* = 3 or 4). **(B)** Calibration curve of HPV-18 detection using SPEEDi-CRISPR shows a wide dynamic range (5 fM - 1 pM) and strong linear relationship (R^2^=0.997), the LoD was 2.3 fM determined by 3σ rule. **(C)** The selectivity of SPEEDi-CRISPR for HPV-18 (10 nM) against HPV 16 (10 nM), PB-19 (10 nM), and scrambled HPV 18 (10 nM). Error bars: one S.D. (*n* = 3 or 4). **(D)** The reproducibility of SPEEDi-CRISPR in five batches of experiment. The RSD among five batches is 2.64%. All measurements were performed by a qPCR device. Error bars: one S.D. (*n*= 3).

For studying the universality of SPEEDi-CRISPR, all tests have been performed for HPV-16 detection (**Fig. S4B, Fig. S5A-C**). An excellent sensitivity (LoD = 4.9 fM, R^2^ = 0.997) and selectivity have been verified for HPV-16 detection, demonstrating that SPEEDi-CRISPR may be utilized to identify a variety of infections. We also tested the reproducibility of SPEEDi-CRISPR, five different batches of the HPV-18 test have been performed, and measurements of fluorescence intensity from each detection were collected as exhibited in **Fig. 4D**. The relative standard deviation (RSD) of signals among different batches is lower than 3%, given that our assay is stable, robust, and repeatable.

### Detection of HPV in spiked samples using the SPEEDi-CRISPR assay

As a proof-of-concept demonstration of potential clinical applications, we adapted the SPEEDi-CRISPR assay to detect both HPV-18 and HPV-16 in spiked samples. Human serum with different spiked HPV concentrations was prepared, 500 μL of each sample was lysed and the DNA was eluted with 100 μL of water as instructed in the DNA extraction kit’s protocol, and the post-purification sample was tested by the SPEEDi-CRISPR assay. HPV DNA spiked in water was used as a control, and the fluorescence intensity of each sample was evaluated and compared to signals from serum samples. As depicted in **Fig. 5A and B**, HPV-18 and HPV-16 DNA isolated from serum and water showed comparable signal intensity at different target DNA concentrations, indicating that SPEEDi-CRISPR has great potential to be applied in complex biological samples. We also evaluated the recovery rate of our assay in the detection of purified DNA from serum, as shown in **Table S2**, the SPEEDi-CRISPR assay can achieve a decent recovery rate ranging from 93.8% to 106% (**Table. S2A**). The HPV DNA isolated from serum samples was also examined by the CRISPR-Dx without extraction. **Fig. S6** shows that routine CRISPR-Dx without extraction is not able to detect the target DNA at a low concentration (200 fM), however, our SPEEDi-CRISPR assay is sensitive enough to generate a strong signal significantly different from the background, exemplifying the enhanced performance of our assay.

**Fig. 5.**
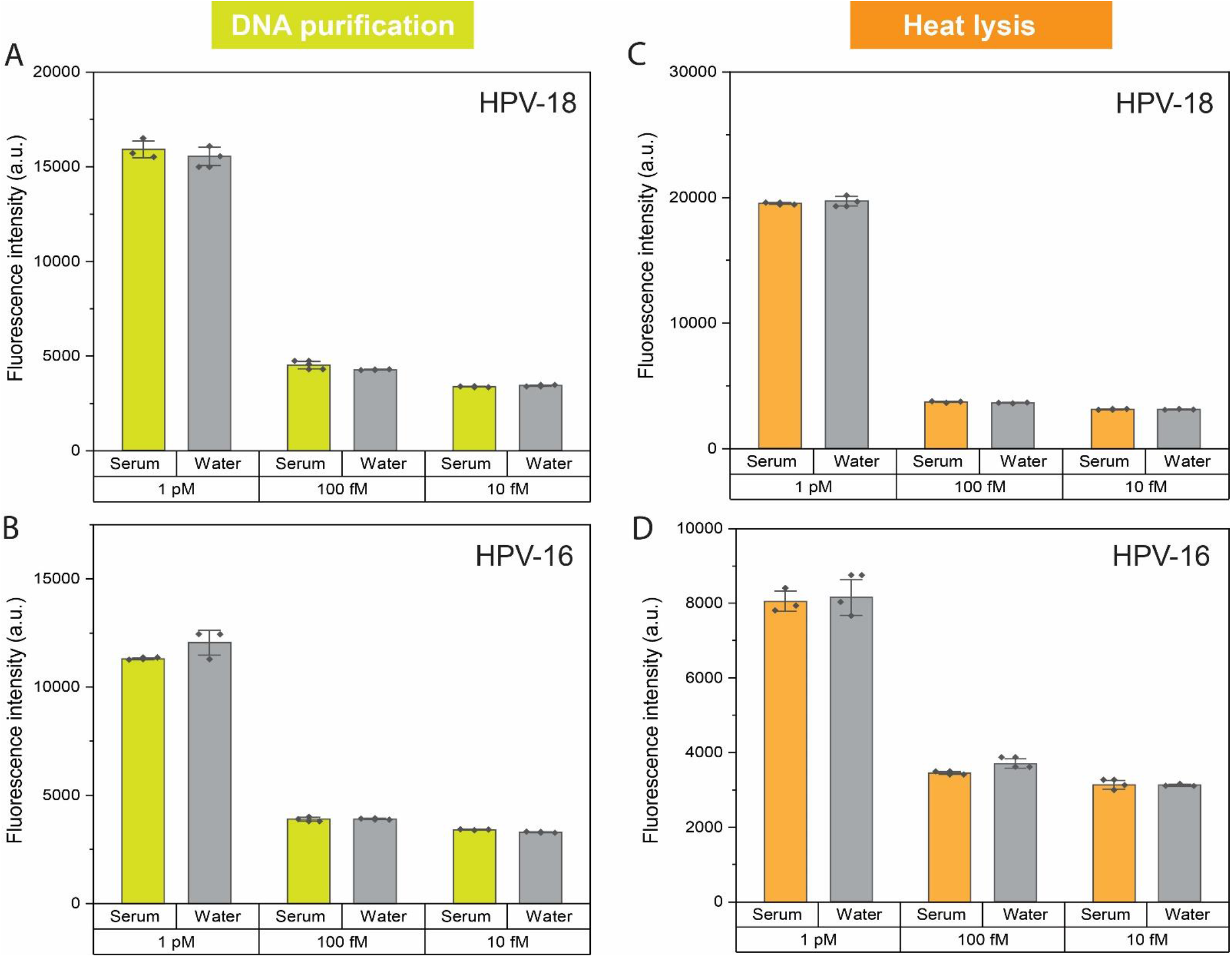
Detection of different concentrations of HPV-18 and HPV-16 in serum samples. **(A)** Detection of HPV-18 in spiked serum samples after DNA purification using the SPEEDi-CRISPR assay. **(B)** Detection of HPV-16 in spiked serum samples after DNA purification using the SPEEDi-CRISPR assay. **(C)** Detection of HPV-18 in spiked serum samples after heat lysis using the SPEEDi-CRISPR assay. **(D)** Detection of HPV-16 in spiked serum samples after heat lysis using the SPEEDi-CRISPR assay. DNA spiked in water was used as a control. All measurements were performed by a qPCR device. Error bars: one S.D. (*n* = 3 or 4).

The column-based DNA purification process used in this work requires multiple reagents and centrifugation. To simplify the detection workflow, thermal lysis that has been developed and demonstrated as a useful lysis method in viral nucleic acid detection^38-41^ was adopted to mimic the virus lysis and HPV DNA release as the sample preparation step. Then the SPEEDi-CRISPR assay was utilized to capture target DNA directly from the serum samples without any DNA purification step. HPV DNA with different concentrations of was spiked into the samples containing 10% human serum and detected by the SPEEDi-CRISPR assay after being heated at 95°C for 10 min, HPV DNA spiked samples containing no serum were used as the control. As shown in **Fig. 5C and D**, HPV-16 and HPV-18 DNA isolated from serum and water showed similar signal intensity at different spiked DNA concentrations, indicating the feasibility of the thermal lysis coupled SPEEDi-CRISPR assay. In addition, our SPEEDi-CRISPR can identify targets in the serum sample after thermal lysis with an excellent recovery rate (89%-103%) and RSD (1.11%-4.34%) (**Table. S2B**). These findings demonstrate that the SPEEDi-CRISPR assay is robust enough to extract DNA directly from the complex biological samples after virus thermal lysis and target DNA release for clinical diagnosis.

### Application of the SPEEDi-CRISPR assay for point-of-care testing

The simplicity and robustness of SPEEDi-CRISPR that we demonstrated above in HPV detection make it intrinsically adaptable to point-of-care (POC) infectious disease diagnostics to promote disease prevention and control in low- and middle-income regions. For this purpose, SPEEDi-CRISPR should utilize an easy and affordable signal readout strategy and be developed into a low-cost, portable POC testing system. With a limited budget, we sought to create a portable assay platform (**Fig. 6A**) that would not merely provide optimum SPEEDi-CRISPR assay conditions but also a low-cost signal readout strategy. Our portable device consists of a consumer digital hotplate (coffee mug warmer) to maintain the reaction’s temperature, a blue LED illuminator, a plastic filter (520 nm long-pass), and 3D-printed body parts. The blue LED is turned on at the end of the test for the detection of fluorescence signals through the filter by a smartphone mounted on a 3D-printed phone holder.

**Fig. 6.**
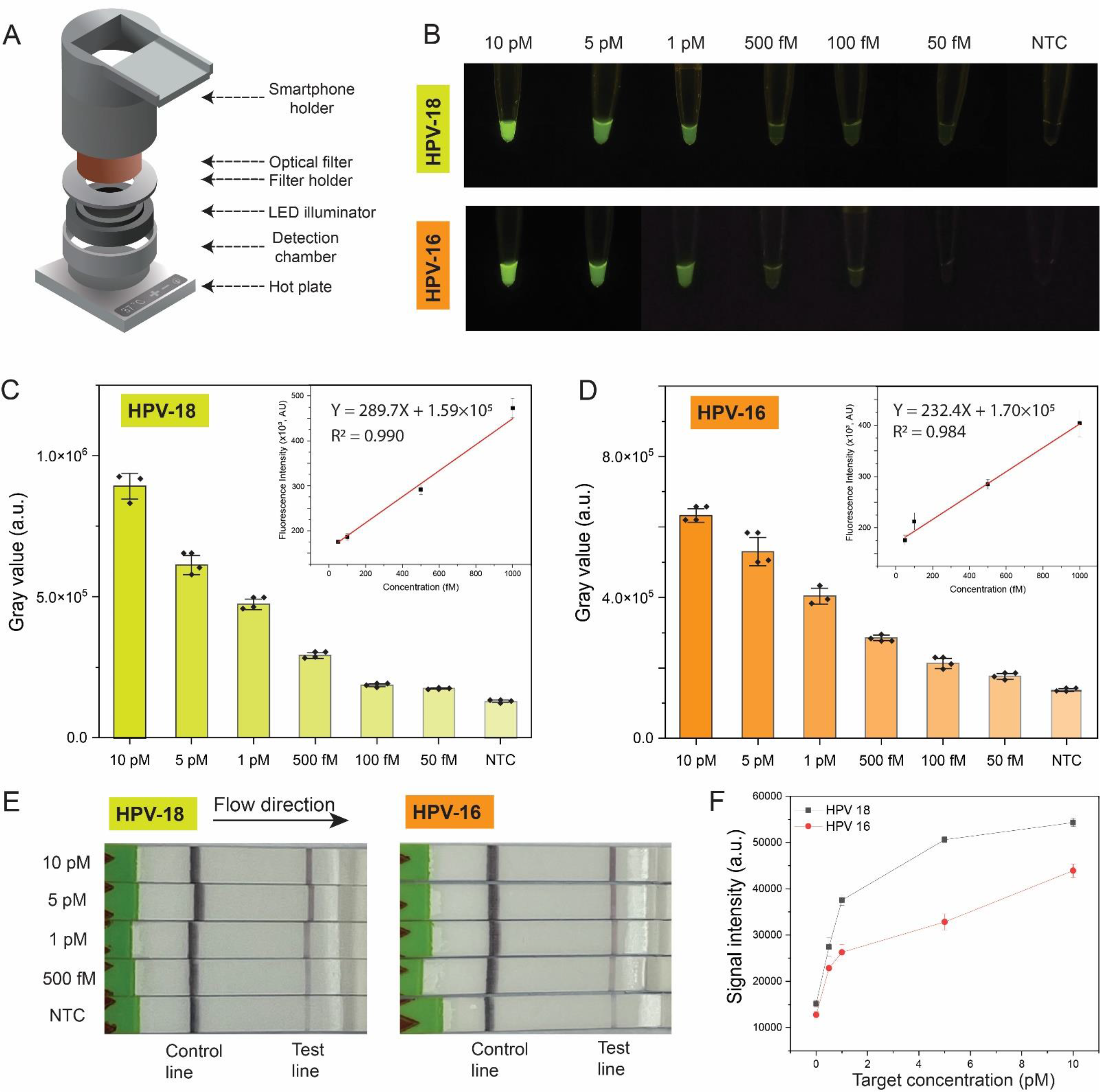
SPEEDi-CRISPR assay for point-of-care testing. **(A)** The exploded view of a portable home-made device assembled with an optical filter, the 3D-printed body parts, a blue LED illuminator, and a consumer digital hotplate. **(B)** Representative fluorescence images of two HPV targets at variable concentrations (10 pM, 5 pM, 1 pM, 500 fM, 100fM, 50 fM, and NTC) captured by the smartphone. **(C-D)** Quantitative analyses and calibration curves for the detection of HPV-18 (C) and HPV-16 (D) using the portable device-coupled SPEEDi-CRISPR assay. The grayscale analysis of the fluorescence signal in each tube is achieved by ImageJ. The LoD was determined by 3σ rule (47.0 fM for HPV-18 and 48.3 fM for HPV-16). Error bars: one S.D. (*n* = 3 or 4). **(E)** Representative photos of LFA assay for two HPV targets at variable concentrations (10 pM, 5 pM, 1 pM, 500 fM, and NTC). **(F)** Measurements of test line band intensity for HPV-18 (black) and HPV-16 (red) by ImageJ. Error bar: one S.D. (*n* = 2).

We first proved the viability of the portable device by conducting the signal generation step of the SPEEDi-CRISPR assay on the hotplate surface. Fluorescence signals from the home-made portable device and the bench-top incubator were compared under identical conditions. **Figure S7** shows that our portable device could preserve almost all the signal intensity of 1 pM HPV-18 compared to that of the incubator, indicating the robustness of the SPEEDi-CRISPR assay and the feasibility of coupling our assay with the home-made portable device. The portable device was then tested for the detection of HPV-16 and HPV-18 at various concentrations (ranging from 10 pM to 50 fM), and we discovered that the SPEEDi-CRISPR assay coupled with the portable device could identify HPV at 50 fM successfully (**Fig. 6B**). The signal intensity of targets with different concentrations has been quantified by ImageJ (**Fig. 6C and D**) and calibrated for analytical performance. Compared to the SPEEDi-CRISPR assay performed in the incubator and the qPCR device (LOD: 2.3 fM), our assay that employed this low-cost portable platform and smartphone still can achieve a LOD of 47.0 fM and 48.3 fM for HPV-18 and HPV-16, respectively. Additionally, the R^2^ of calibration curves for the detection of HPV-18 and HPV-16 in the dynamic range (50 fM – 1 pM) is 0.990 and 0.984, respectively, indicating the quantitative potential of our assay for the accurate POC testing of HPV.

Lateral flow assay (LFA) is another broadly used method in POC testing because it eliminates the demand of specialized and costly equipment and personnel. To further demonstrate the potential POC applications of our assay, we attempted to combine the SPEEDi-CRISPR assay with a commercially available LFA test strip for instrument-free readout. In this case, the SPEEDi-CRISPR assay was performed by replacing the FQ-ssDNA reporter with a FAM-Biotin ssDNA (FB-ssDNA) reporter in the reaction buffer (**Fig. S8**). The FB-ssDNA reporters bound on the immunogold nanoparticles can be cleaved by the target-activated RNPs and the cleaved immunogold-reporter complexes can be captured by the secondary antibody deposited on the test line on the strip to yield the positive signal.^25^ Finally, signals for various HPV DNA concentrations were evaluated with lateral flow dipsticks (10 pM-500 fM). The LFA-based SPEEDi-CRISPR assay, as shown in **Fig. 6E**, can clearly distinguish HPV samples (10 pM, 5 pM, 1 pM, and 500 fM) from negative samples, and higher concentrations of HPV reflect a stronger intensity of test lines. Compared to the fluorescence detection approach (**Fig. 6C&D**), visual LFA detection displayed inferior performance for quantitative analysis with decreased linearity and dynamic range (**Fig. 6F**). However, LFA-coupled SPEEDi-CRISPR assay can still be used for the rapid and convenient detection of HPV as a qualitative or semiquantitative method in resource-limited regions.

## Discussion

The high specificity, programmability, and ease of use of CRISPR technology make it the next-generation diagnostic platform. A lot of CRISPR-based biosensing tools^20-22, 42-44^ and materials^45^,^46^ have been developed based on the collateral cleavage (trans cleavage) activity of Cas12a. Though the trans-cleavage ability of Cas12a enables signal amplification in diagnostics, the limited sensitivity still restricts the application of Cas12a in amplification-free diagnosis. To overcome this issue, we developed an assay termed SPEEDi-CRISPR for solid-phase extraction and enhanced detection of pathogens. The SPEEDi-CRISPR assay harnesses the specific target sequence recognition and indiscriminate trans-cleavage ability of Cas12a simultaneously. Cas12a-crRNA RNP is modified on magnetic beads and its target recognition ability is used in solid-phase extraction for the first time, after target enrichment, the beads can be utilized for subsequent signal generation. As a result, the dual functions of Cas12a integrated into a single solid-phase extraction material in our assay afford the sensitive, rapid, and convenient detection of pathogens in an amplification-free way.

Cervical cancer is the fourth most common cancer in women worldwide, with more than 600,000 new cases and more than 300,000 deaths in 2020. About 90% of the new cases and deaths worldwide in 2020 took place in low- and middle-income countries, and the human papillomavirus (HPV) accounts for more than 95% of cervical cancer cases.^47^ Therefore, a rapid, sensitive, and cost- and reagent-effective screening of HPV will dramatically benefit the diagnosis and treatment, especially in resource-limited areas. In this work, we used HPV-18 and HPV-16, the two high-risk HPV subtypes, as models to evaluate the performance of our SPEEDi-CRISPR assay. Compared to the other CRISPR-Dx (**Table S3**), our assay exhibits high sensitivity and outstanding simplicity with a detection limit of 2.3 fM for HPV-18 and 4.9 fM for HPV-16 with a wide linear range from 1 pM to 5 fM. Our isothermal amplification-free assay can be completed in 2 hours and can dramatically simplify the workflow and reduce the dependence on precise laboratory equipment. The SPEEDi-CRISPR assay was robust enough for HPV detection in spiked serum samples, indicating the great potential in the analysis of complicated biological samples. The great adaptability of the SPEEDi-CRISPRT also enables the coupling with a smartphone-based portable device and LFA, proving that our assay is adequate for POC testing. Altogether, we envision this new technology holds great promise for pathogen diagnosis as an infection control strategy globally, particularly in nations with low resources.

## Methods

RNA and DNA oligos (**Table. S1**) were purchased from IDT (Integrated DNA Technologies). Dynabeads™ His-Tag Isolation & Pulldown was purchased from Invitrogen Corporation (Carlsbad, California, USA). NEBuffer™ 2.1 from New England Biolabs Ltd. (New England, USA) is selected as the buffer to control the pH value and provide an enzyme-friendly environment. UltraPureTM Distilled water which is DNase and RNase free is purchased from Invitrogen Corporation (Carlsbad, California, USA). EnGen® Lba Cas12a (Cpf1) from New England Biolabs Ltd. (New England, USA) is selected in this work. During the reaction, the Eppendorf thermomixer C was used for incubation, and the CFX Connect Real-Time PCR Detection System (Bio-Rad) is used to detect fluorescence intensity.

### Performing Cas12a’s trans-cleavage activity in free solution

The RNP is prepared by incubating 200 nM Cas12a and 300 nM crRNA in 1 × NE buffer 2.1 at 37 °C for 30 min. The target double-strand DNA is annealed with the ratio of 2:1 (2 parts of complementary strand and 1 part of target strand) after heating to 95°C for 5 min. The trans-cleavage reaction is performed with 50 nM RNP, 50 nM FQ-ssDNA reporter and 1 x NE buffer with different target dsDNA concentrations in 200 μL tube strips at 37°C for 120 min, and the fluorescence intensity is monitored every 1 min. The final concentration of target double-strand DNA in the 20-μL reaction system is used to determine the analytical sensitivity.

### Preparation of the RNPs-coated beads

The RNP is prepared by incubating 200 nM LbuCas12a which has a N-terminal 6×his-tagged with 300 nM crRNA in 1 × NE buffer 2.1 at 37 °C for 30 min. Under the condition of 0.02% Tween™-20, incubate the prepared RNP with Dynabeads™ His-Tag Isolation & Pulldown at room temperature for 10 min. After incubating, the prepared beads are thoroughly rinsed with wash buffer which contains 1 × NE buffer 2.1 and 0.02% Tween™-20, and stored at 4 °C for ready.

### Measurement of the zeta potential of RNPs-coated beads

The zeta potential of fresh preparation beads was determined by ZetaView system (Particle Metrix Inc.). RNPs-coated beads were diluted in 1x PBS and 0.02% Tween-20 to an optimal concentration according to the manufacturer’s recommendations.

### Exploring the analytical performance of SPEEDi-CRISPR

SPEEDi-CRISPR assay contains the target enrichment step and the signal generation step. In the enrichment step, RNPs-coated beads were added into the sample (100 μL) that contains target DNA, 1 × NE buffer 2.1, and 0.02% Tween™-20 and incubated at 37 °C. After enrichment, the supernatant is discarded, and post-enrichment RNPs-coated beads were added into the reaction buffer (20 μL) with 600 nM FQ-ssDNA reporter, 1 × NE buffer 2.1 and 0.02% Tween™-20 and incubated at 37 °C for 80 min, and the end-time fluorescence intensity is measured. The final concentration of target DNA from the same sample in the 20 μL free-solution Cas12a reaction system is used to determine the analytical sensitivity.

### Spike serum sample for biosensor assay

Spike serum samples were purified by DNA Isolation Kit - Plasma/Serum (ab156893). Elution samples (100 μL) were detected by SPEEDi-CRISPR under optimal conditions.

### Quantification in heat-lysis serum samples with the SPEEDi-CRISPR assay

The biological serum samples were obtained from Innovative Research (MI, USA). 10 μL of serum is added to 90 μL beads wash buffer, and the solution was heated to 95°C for 10 min to lyse samples and deactivate deoxyribonucleases. Finally, different concentrations of target DNA were added to the serum sample and were detected by SPEEDi-CRISPR.

### SPEEDi-CRISPR performed by portable device

A customer digital hotplate and a 450 nm LED illuminator were purchased from Amazon and Boli Optics, respectively, and were assembled with a scaffolding that was designed using AutoCAD 2022 and fabricated by 3D printing (FormLabs Form 3+ SLA 3D printer). To achieve our assay in the portable device, hotplate heating at 37 °C was used for both enrichment and reaction. Furthermore, shaking tubes by hand every 30 min could make the performance as good as that achieved by an incubator. Finally, the gray value of each reaction was analyzed by ImageJ software.

### SPEEDi-CRISPR performed by LFA

The LFA kit was purchased from Milenia Biotec (HybriDetect – Universal Lateral Flow Assay Kit). The lateral flow assay is using activated RNPs-coated beads to cleave the FAM-biotin labeled ssDNA reporter (Biotin-TTATT-FAM, IDT). After the reaction, 80 μL of HybriDetect assay buffer was added to the 20-μL reaction supernatant and incubated at room temperature for 5 min. Milena HybriDetect1 lateral flow dipstick (TwistDx) is dipped into the mixture and incubated for 5 minutes at room temperature. Then, the dipstick is removed, and the results are interpreted immediately by photograph using a smartphone camera and analyzed by Image J. A negative control is an assay without target DNA and should have an intense line at the control line. Finally, the gray value of each reaction was analyzed by ImageJ software.

## Data Availability

All data produced in the present work are contained in the manuscript

## ASSOCIATED CONTENT

### Supporting information

Additional information and results (Fig. S1-S8 and Tables S1-S3).

## AUTHOR INFORMATION

### Author contributions

Z.T. performed experiments. Z.T., H.Y., and Y.Z. interpreted the data. H.Y., and Y.Z. supervised the work. Z.T., H.Y., and Y.Z. made substantial contributions to the conception and design of the work. Z.T., H.Y., and Y.Z. contributed to the draft of the work.

## ACKNOWLEDGMENT

This study was supported in part by the grants R01 CA243445, R33 CA214333, R33CA252158A1, and R01CA260132 from National Institutes of Health.

## REFERENCE

1. Chen, Y.; Qian, C.; Liu, C.; Shen, H.; Wang, Z.; Ping, J.; Wu, J.; Chen, H., Nucleic acid amplification free biosensors for pathogen detection. Biosens. Bioelectron. 2020, 153, 112049.

2. Dai, Y.; Liu, C. C., Recent Advances on Electrochemical Biosensing Strategies toward Universal Point-of-Care Systems. Angew. Chem., Int. Ed. Engl. 2019, 58 (36), 12355–12368.

3. McConnell, E. M.; Cozma, I.; Morrison, D.; Li, Y., Biosensors Made of Synthetic Functional Nucleic Acids Toward Better Human Health. Anal. Chem. 2020, 92 (1), 327–344.

4. Smith, S. J.; Nemr, C. R.; Kelley, S. O., Chemistry-Driven Approaches for Ultrasensitive Nucleic Acid Detection. J. Am. Chem. Soc. 2017, 139 (3), 1020–1028.

5. Song, P.; Li, M.; Shen, J.; Pei, H.; Chao, J.; Su, S.; Aldalbahi, A.; Wang, L.; Shi, J.; Song, S.; Wang, L.; Fan, C.; Zuo, X., Dynamic Modulation of DNA Hybridization Using Allosteric DNA Tetrahedral Nanostructures. Anal. Chem. 2016, 88 (16), 8043–9.

6. Zhou, H.; Liu, J.; Xu, J. J.; Zhang, S. S.; Chen, H. Y., Optical nano-biosensing interface via nucleic acid amplification strategy: construction and application. Chem. Soc. Rev. 2018, 47 (6), 1996–2019.

7. Chen, J. S.; Ma, E.; Harrington, L. B.; Da Costa, M.; Tian, X.; Palefsky, J. M.; Doudna, J. A., CRISPR-Cas12a target binding unleashes indiscriminate single-stranded DNase activity. Science 2018, 360 (6387), 436–439.

8. Joung, J.; Ladha, A.; Saito, M.; Kim, N. G.; Woolley, A. E.; Segel, M.; Barretto, R. P. J.; Ranu, A.; Macrae, R. K.; Faure, G.; Ioannidi, E. I.; Krajeski, R. N.; Bruneau, R.; Huang, M. W.; Yu, X. G.; Li, J. Z.; Walker, B. D.; Hung, D. T.; Greninger, A. L.; Jerome, K. R.; Gootenberg, J. S.; Abudayyeh, O. O.; Zhang, F., Detection of SARS-CoV-2 with SHERLOCK One-Pot Testing. N. Engl. J. Med. 2020, 383 (15), 1492–1494.

9. Kellner, M. J.; Koob, J. G.; Gootenberg, J. S.; Abudayyeh, O. O.; Zhang, F., SHERLOCK: nucleic acid detection with CRISPR nucleases. Nat. Protoc. 2019, 14 (10), 2986–3012.

10. Konwarh, R., Can CRISPR/Cas Technology Be a Felicitous Stratagem Against the COVID-19 Fiasco? Prospects and Hitches. Front Mol. Biosci. 2020, 7, 557377.

11. Patchsung, M.; Jantarug, K.; Pattama, A.; Aphicho, K.; Suraritdechachai, S.; Meesawat, P.; Sappakhaw, K.; Leelahakorn, N.; Ruenkam, T.; Wongsatit, T.; Athipanyasilp, N.; Eiamthong, B.; Lakkanasirorat, B.; Phoodokmai, T.; Niljianskul, N.; Pakotiprapha, D.; Chanarat, S.; Homchan, A.; Tinikul, R.; Kamutira, P.; Phiwkaow, K.; Soithongcharoen, S.; Kantiwiriyawanitch, C.; Pongsupasa, V.; Trisrivirat, D.; Jaroensuk, J.; Wongnate, T.; Maenpuen, S.; Chaiyen, P.; Kamnerdnakta, S.; Swangsri, J.; Chuthapisith, S.; Sirivatanauksorn, Y.; Chaimayo, C.; Sutthent, R.; Kantakamalakul, W.; Joung, J.; Ladha, A.; Jin, X.; Gootenberg, J. S.; Abudayyeh, O. O.; Zhang, F.; Horthongkham, N.; Uttamapinant, C., Clinical validation of a Cas13-based assay for the detection of SARS-CoV-2 RNA. Nat. Biomed. Eng. 2020, 4 (12), 1140–1149.

12. Kaminski, M. M.; Abudayyeh, O. O.; Gootenberg, J. S.; Zhang, F.; Collins, J. J., CRISPR-based diagnostics. Nat. Biomed. Eng. 2021, 5 (7), 643–656.

13. Aman, R.; Mahas, A.; Mahfouz, M., Nucleic Acid Detection Using CRISPR/Cas Biosensing Technologies. ACS Synth. Biol. 2020, 9 (6), 1226–1233.

14. Chen, B.; Li, Y.; Xu, F.; Yang, X., Powerful CRISPR-Based Biosensing Techniques and Their Integration With Microfluidic Platforms. Front. Bioeng. Biotechnol. 2022, 10, 851712.

15. Li, Y.; Li, S.; Wang, J.; Liu, G., CRISPR/Cas systems towards next-generation biosensing. Trends Biotechnol. 2019, 37 (7), 730–743.

16. East-Seletsky, A.; O’Connell, M. R.; Knight, S. C.; Burstein, D.; Cate, J. H.; Tjian, R.; Doudna, J. A., Two distinct RNase activities of CRISPR-C2c2 enable guide-RNA processing and RNA detection. Nature 2016, 538 (7624), 270–273.

17. Li, S. Y.; Cheng, Q. X.; Liu, J. K.; Nie, X. Q.; Zhao, G. P.; Wang, J., CRISPR-Cas12a has both cis- and trans-cleavage activities on single-stranded DNA. Cell Res. 2018, 28 (4), 491–493.

18. Peng, S.; Tan, Z.; Chen, S.; Lei, C.; Nie, Z., Integrating CRISPR-Cas12a with a DNA circuit as a generic sensing platform for amplified detection of microRNA. Chem. Sci. 2020, 11 (28), 7362–7368.

19. Xiong, Y.; Zhang, J.; Yang, Z.; Mou, Q.; Ma, Y.; Xiong, Y.; Lu, Y., Functional DNA Regulated CRISPR-Cas12a Sensors for Point-of-Care Diagnostics of Non-Nucleic-Acid Targets. J. Am. Chem. Soc. 2020, 142 (1), 207–213.

20. Bruch, R.; Baaske, J.; Chatelle, C.; Meirich, M.; Madlener, S.; Weber, W.; Dincer, C.; Urban, G. A., CRISPR/Cas13a-Powered Electrochemical Microfluidic Biosensor for Nucleic Acid Amplification-Free miRNA Diagnostics. Adv. Mater. 2019, 31 (51), e1905311.

21. Dai, Y.; Somoza, R. A.; Wang, L.; Welter, J. F.; Li, Y.; Caplan, A. I.; Liu, C. C., Exploring the Trans-Cleavage Activity of CRISPR-Cas12a (cpf1) for the Development of a Universal Electrochemical Biosensor. Angew. Chem. Int. Ed. Engl. 2019, 58 (48), 17399–17405.

22. Han, C.; Li, W.; Li, Q.; Xing, W.; Luo, H.; Ji, H.; Fang, X.; Luo, Z.; Zhang, L., CRISPR/Cas12a-Derived electrochemical aptasensor for ultrasensitive detection of COVID-19 nucleocapsid protein. Biosens. Bioelectron. 2022, 200, 113922.

23. Broughton, J. P.; Deng, X.; Yu, G.; Fasching, C. L.; Servellita, V.; Singh, J.; Miao, X.; Streithorst, J. A.; Granados, A.; Sotomayor-Gonzalez, A.; Zorn, K.; Gopez, A.; Hsu, E.; Gu, W.; Miller, S.; Pan, C. Y.; Guevara, H.; Wadford, D. A.; Chen, J. S.; Chiu, C. Y., CRISPR-Cas12-based detection of SARS-CoV-2. Nat. Biotechnol. 2020, 38 (7), 870–874.

24. Qian, C.; Wang, R.; Wu, H.; Zhang, F.; Wu, J.; Wang, L., Uracil-Mediated New Photospacer-Adjacent Motif of Cas12a To Realize Visualized DNA Detection at the Single-Copy Level Free from Contamination. Anal. Chem. 2019, 91 (17), 11362–11366.

25. Gootenberg, J. S.; Abudayyeh, O. O.; Kellner, M. J.; Joung, J.; Collins, J. J.; Zhang, F., Multiplexed and portable nucleic acid detection platform with Cas13, Cas12a, and Csm6. Science 2018, 360 (6387), 439–444.

26. Wang, B.; Wang, R.; Wang, D.; Wu, J.; Li, J.; Wang, J.; Liu, H.; Wang, Y., Cas12aVDet: A CRISPR/Cas12a-Based Platform for Rapid and Visual Nucleic Acid Detection. Anal. Chem. 2019, 91 (19), 12156–12161.

27. Lippi, G.; von Meyer, A.; Cadamuro, J.; Simundic, A. M., Blood sample quality. Diagnosis (Berl) 2019, 6 (1), 25–31.

28. Sanchez-Giron, F.; Alvarez-Mora, F., Reduction of blood loss from laboratory testing in hospitalized adult patients using small-volume (pediatric) tubes. Arch. Pathol. Lab. Med. 2008, 132 (12), 1916–9.

29. Wisser, D.; van Ackern, K.; Knoll, E.; Wisser, H.; Bertsch, T., Blood loss from laboratory tests. Clin. Chem. 2003, 49 (10), 1651–5.

30. Pulford, D. J.; Mosteller, M.; Briley, J. D.; Johansson, K. W.; Nelsen, A. J., Saliva sampling in global clinical studies: the impact of low sampling volume on performance of DNA in downstream genotyping experiments. BMC Med. Genomics 2013, 6, 20.

31. Callahan, C.; Lee, R. A.; Lee, G. R.; Zulauf, K.; Kirby, J. E.; Arnaout, R., Nasal Swab Performance by Collection Timing, Procedure, and Method of Transport for Patients with SARS-CoV-2. J. Clin. Microbiol. 2021, 59 (9), e0056921.

32. Crosbie, E. J.; Einstein, M. H.; Franceschi, S.; Kitchener, H. C., Human papillomavirus and cervical cancer. Lancet 2013, 382 (9895), 889–99.

33. Schiffman, M.; Castle, P. E.; Jeronimo, J.; Rodriguez, A. C.; Wacholder, S., Human papillomavirus and cervical cancer. Lancet 2007, 370 (9590), 890–907.

34. Nalefski, E. A.; Patel, N.; Leung, P. J. Y.; Islam, Z.; Kooistra, R. M.; Parikh, I.; Marion, E.; Knott, G. J.; Doudna, J. A.; Le Ny, A. M.; Madan, D., Kinetic analysis of Cas12a and Cas13a RNA-Guided nucleases for development of improved CRISPR-Based diagnostics. iScience 2021, 24 (9), 102996.

35. Kim, H.; Lee, W. J.; Oh, Y.; Kang, S. H.; Hur, J. K.; Lee, H.; Song, W.; Lim, K. S.; Park, Y. H.; Song, B. S.; Jin, Y. B.; Jun, B. H.; Jung, C.; Lee, D. S.; Kim, S. U.; Lee, S. H., Enhancement of target specificity of CRISPR-Cas12a by using a chimeric DNA-RNA guide. Nucleic Acids Res. 2020, 48 (15), 8601–8616.

36. Strohkendl, I.; Saifuddin, F. A.; Rybarski, J. R.; Finkelstein, I. J.; Russell, R., Kinetic Basis for DNA Target Specificity of CRISPR-Cas12a. Mol. Cell 2018, 71 (5), 816–824.e3.

37. Singh, D.; Mallon, J.; Poddar, A.; Wang, Y.; Tippana, R.; Yang, O.; Bailey, S.; Ha, T., Real-time observation of DNA target interrogation and product release by the RNA-guided endonuclease CRISPR Cpf1 (Cas12a). Proc. Natl. Acad. Sci. U S A 2018, 115 (21), 5444–5449.

38. Leta, D.; Gutema, G.; Hagos, G. G.; Diriba, R.; Bulti, G.; Sura, T.; Ayana, D.; Chala, D.; Lenjiso, B.; Bulti, J.; Abdella, S.; Tola, H. H., Effect of heat inactivation and bulk lysis on real-time reverse transcription PCR detection of the SARS-COV-2: an experimental study. BMC Res. Notes 2022, 15 (1), 295.

39. Qian, J.; Boswell, S. A.; Chidley, C.; Lu, Z. X.; Pettit, M. E.; Gaudio, B. L.; Fajnzylber, J. M.; Ingram, R. T.; Ward, R. H.; Li, J. Z.; Springer, M., An enhanced isothermal amplification assay for viral detection. Nat. Commun. 2020, 11 (1), 5920.

40. Steinau, M.; Patel, S. S.; Unger, E. R., Efficient DNA extraction for HPV genotyping in formalin-fixed, paraffin-embedded tissues. J. Mol. Diagn. 2011, 13 (4), 377–81.

41. Thom, R. E.; Eastaugh, L. S.; O’Brien, L. M.; Ulaeto, D. O.; Findlay, J. S.; Smither, S. J.; Phelps, A. L.; Stapleton, H. L.; Hamblin, K. A.; Weller, S. A., Evaluation of the SARS-CoV-2 Inactivation Efficacy Associated With Buffers From Three Kits Used on High-Throughput RNA Extraction Platforms. Front. Cell. Infect. Microbiol. 2021, 11, 716436.

42. Suea-Ngam, A.; Howes, P. D.; deMello, A. J., An amplification-free ultra-sensitive electrochemical CRISPR/Cas biosensor for drug-resistant bacteria detection. Chem. Sci. 2021, 12 (38), 12733–12743.

43. Li, Y.; Mansour, H.; Watson, C. J. F.; Tang, Y.; MacNeil, A. J.; Li, F., Amplified detection of nucleic acids and proteins using an isothermal proximity CRISPR Cas12a assay. Chem. Sci. 2021, 12 (6), 2133–2137.

44. Nouri, R.; Jiang, Y.; Lian, X. L.; Guan, W., Sequence-Specific Recognition of HIV-1 DNA with Solid-State CRISPR-Cas12a-Assisted Nanopores (SCAN). ACS Sens. 2020, 5 (5), 1273–1280.

45. Fu, X.; Shi, Y.; Peng, F.; Zhou, M.; Yin, Y.; Tan, Y.; Chen, M.; Yin, X.; Ke, G.; Zhang, X. B., Exploring the Trans-Cleavage Activity of CRISPR/Cas12a on Gold Nanoparticles for Stable and Sensitive Biosensing. Anal. Chem. 2021, 93 (11), 4967–4974.

46. English, M. A.; Soenksen, L. R.; Gayet, R. V.; de Puig, H.; Angenent-Mari, N. M.; Mao, A. S.; Nguyen, P. Q.; Collins, J. J., Programmable CRISPR-responsive smart materials. Science 2019, 365 (6455), 780–785.

47. Sung, H.; Ferlay, J.; Siegel, R. L.; Laversanne, M.; Soerjomataram, I.; Jemal, A.; Bray, F., Global Cancer Statistics 020: GLOBOCAN Estimates of Incidence and Mortality Worldwide for 36 Cancers in 185 Countries. CA Cancer J. Clin. 2021, 71 (3), 209–249.

