## Supplemental information for "Solid-Phase Extraction and Enhanced Amplification-Free Detection of Pathogens Integrated by Dual-Functional CRISPR-Cas12a"

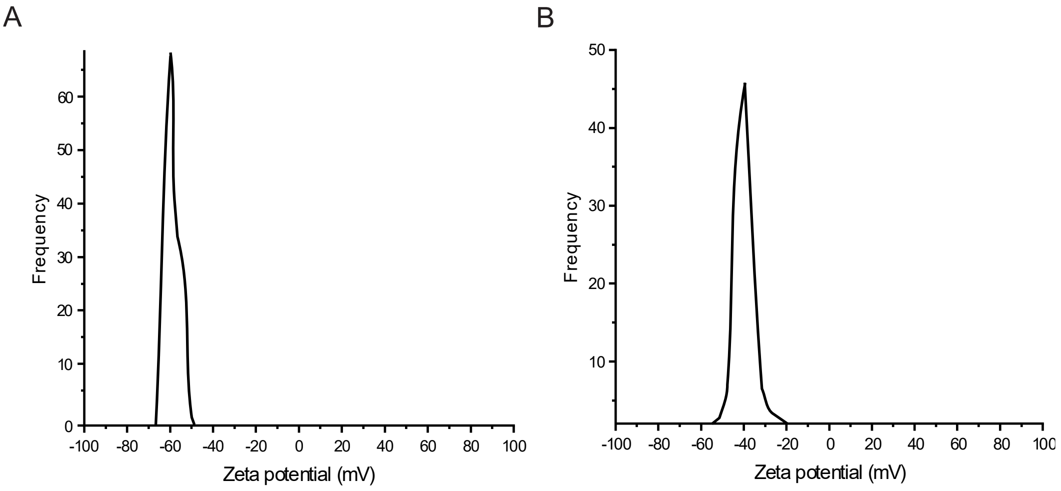


**Figure S1. Zeta potential of RNP-coated beads and uncoated beads. (A)** The zeta potential of RNP-coated beads. **(B)** The zeta potential of uncoated dynabeads. All results were obtained by ZetaView in 1X PBS.

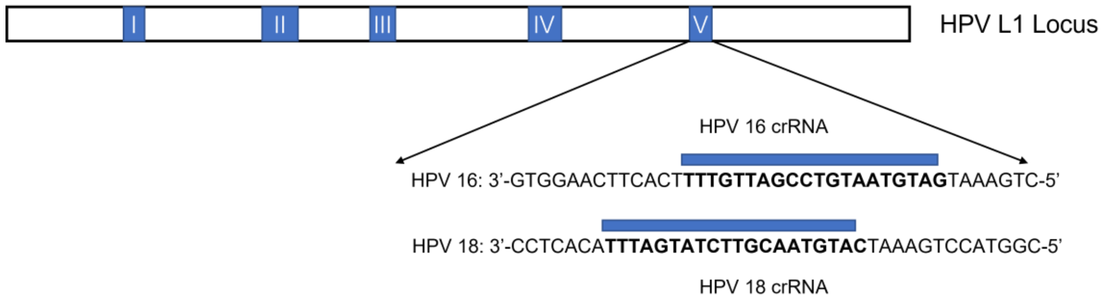


**Fig. S2. Diagram of HPV16 and HPV18 sequences within the hypervariable loop V of the L1-encoding gene targeted by Cas12a.** The bold letters indicate the target sequences of crRNAs.

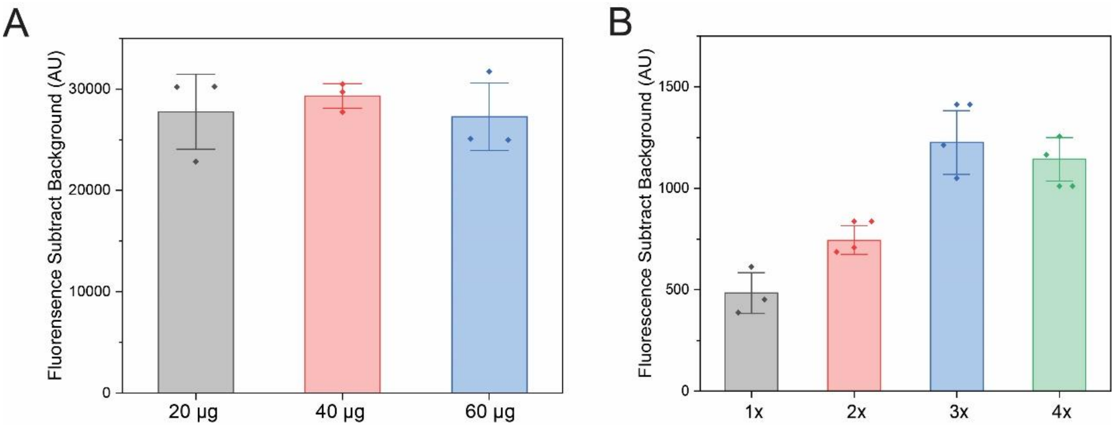


**Fig. S3. Optimization of the SPEEDi-CRISPR assay. (A)** Fluorescence intensities for various amounts of RNPs-coated beads utilized in the SPEEDi-CRISPR experiment. The results indicated that the signal intensity was unaffected by increasing the amount of RNPs-coated beads in the reaction from 20 µg to 60 µg. **(B)** Fluorescence intensity for different rounds of extraction in the assay. The results indicated that the signal intensity would increase until it reached three rounds of extraction and then started to decrease after four rounds. All measurements were performed by a qPCR device. Error bars: one S.D. ($n$ = 3 or 4).

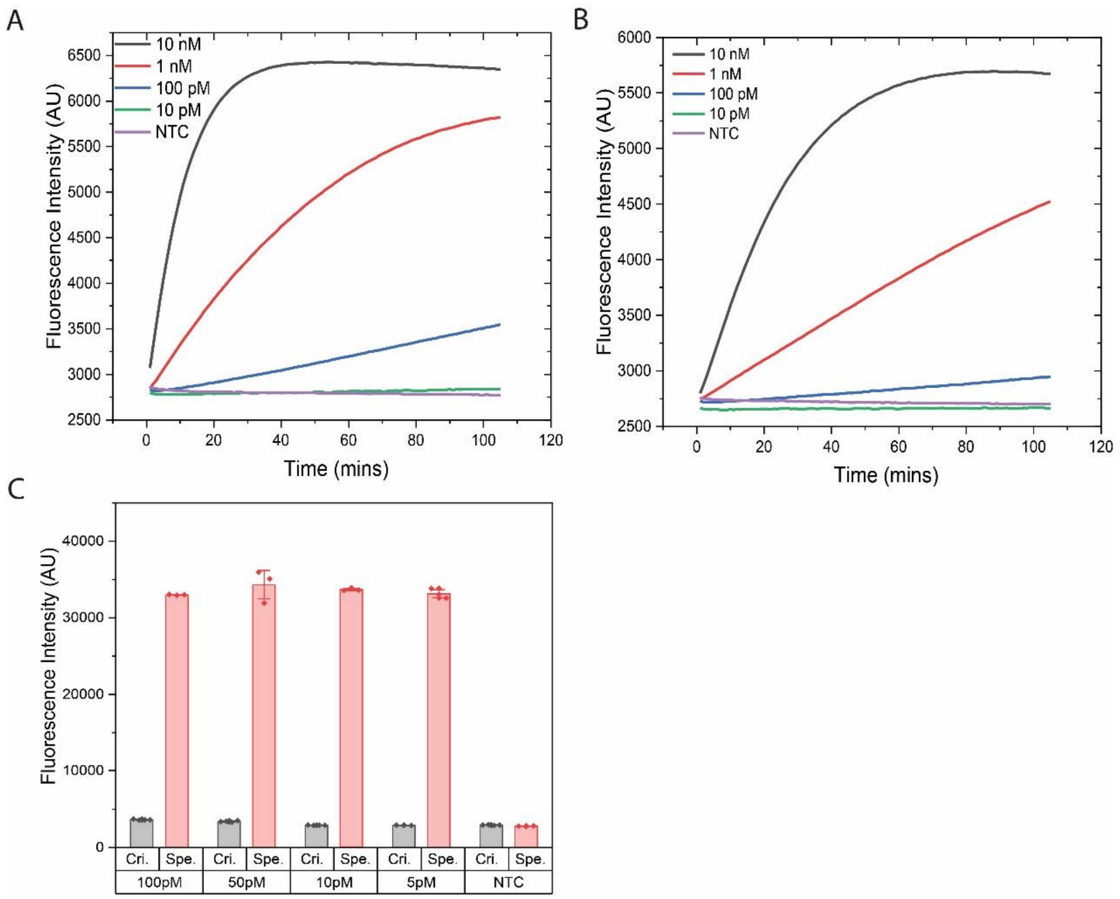


**Fig. S4. Analysis of the analytical performance of traditional CRISPR-Dx. (A)** Real-time fluorescence intensity of Cas12a trans-cleavage reaction with different HPB-18 target concentrations. **(B)** Real-time fluorescence intensity of Cas12a trans-cleavage reaction with different HPB-16 target concentrations. All measurements were performed by the qPCR device. **(C)** Evaluation of the CRISPR/Cas12a assay in free solution without extraction (Cri.) and SPEEDi-CRISPR (Spe.) signal intensities at various target HPV-18 DNA concentrations. All measurements were performed by a qPCR device. Error bars: one S.D. ($n$ = 3 or 4).

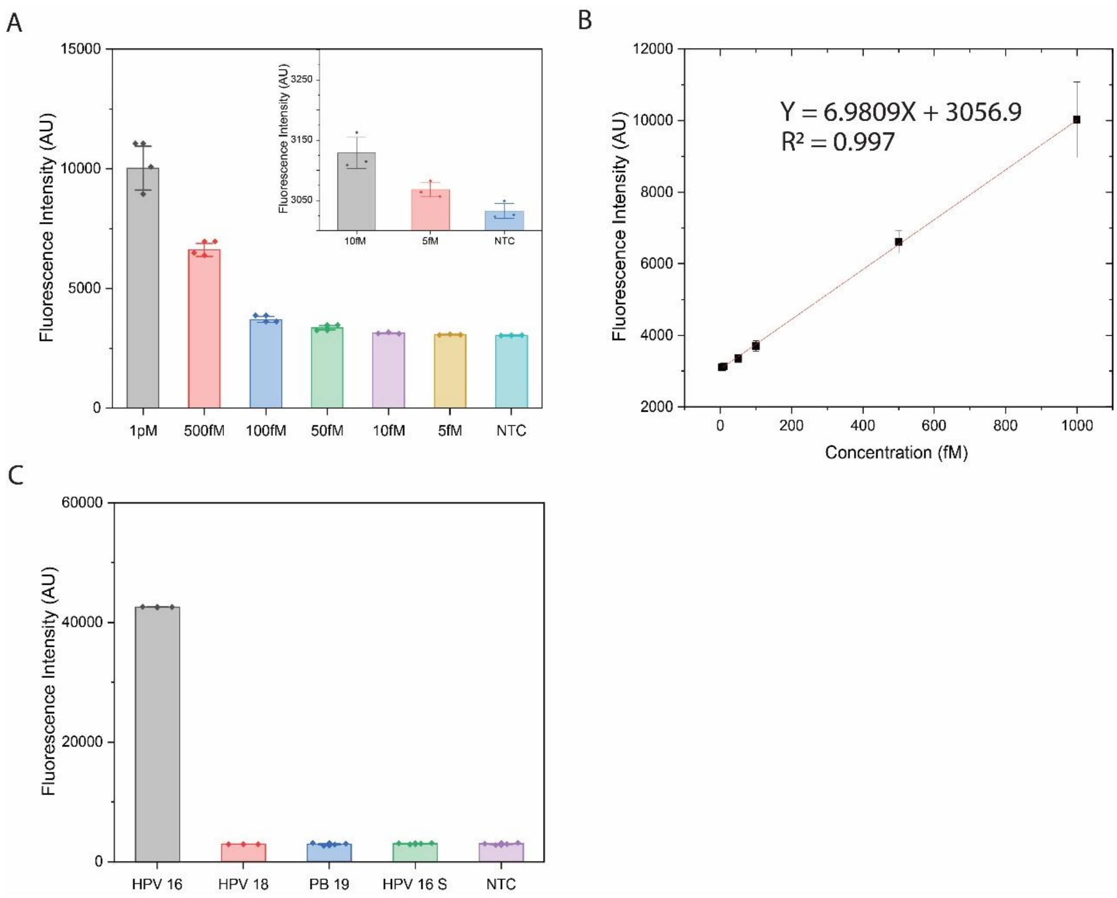


**Fig. S5. Analytical performance of SPEEDi-CRISPR in the detection of HPV-16. (A)** Fluorescence intensity of experiments with different target concentrations in optimal conditions. **(B)** Calibration curve of HPV16 detection using SPEEDi-CRISPR shows a wide dynamic range (5 fM - 1 pM) and strong linear relationship ($R^2$=0.997), the LoD was determined by 3σ rule (4.9 fM). **(C)** The selectivity of SPEEDi-CRISPR for HPV 16 (10 nM) against HPV 18 (10 nM), PB-19 (10 nM), and scrambled HPV 16 (10 nM). All measurements were performed by a qPCR device. Error bars: one S.D. (n= 3).

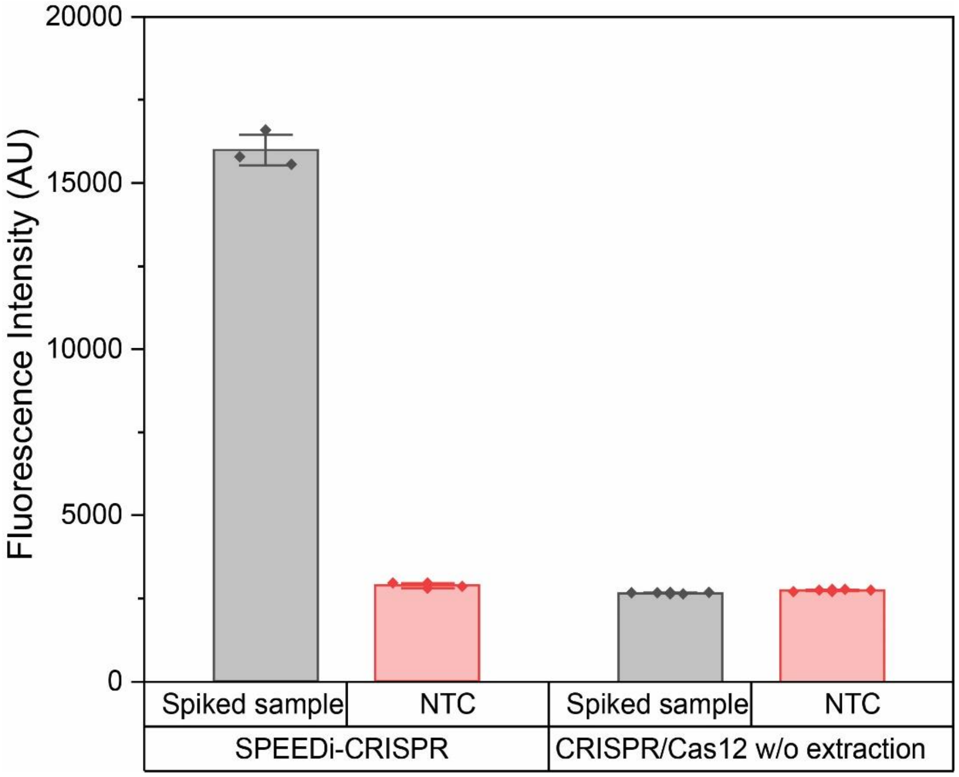


**Fig. S6. Performance comparison of the SPEEDi-CRISPR assay to CRISPR/Cas12a assay without extraction.** The significant difference in signal intensity indicated that the SPEEDi-CRISPR assay can successfully detect low-concentration targets in real samples (200 fM) whereas the CRISPR/Cas12a assay without extraction failed. All measurements were performed by a qPCR device. Error bars: one S.D. (*n* = 3 or 4).

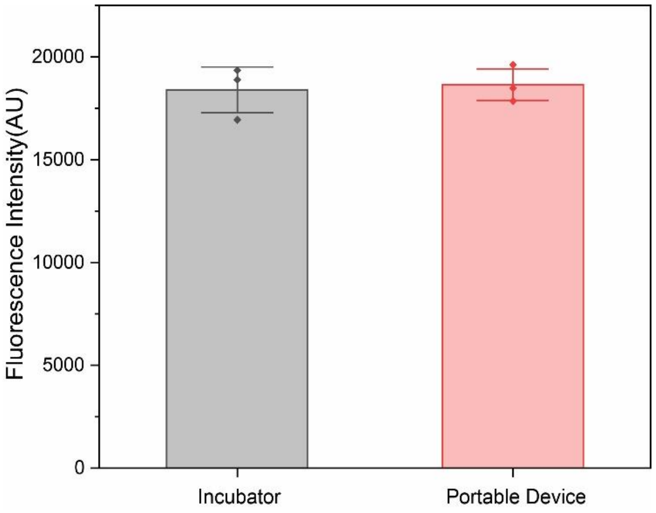


**Fig. S7. Fluorescence intensity of assays in the incubator and the portable device for the detection of 1 pM HPV-18.** Our portable device could preserve ~102% of the fluorescence intensity obtained with a bench-top incubator. The result demonstrated the viability of the POCT method and the robustness of the assay. All measurements were performed by plate-reader, error bars: one S.D. (*n* = 3).

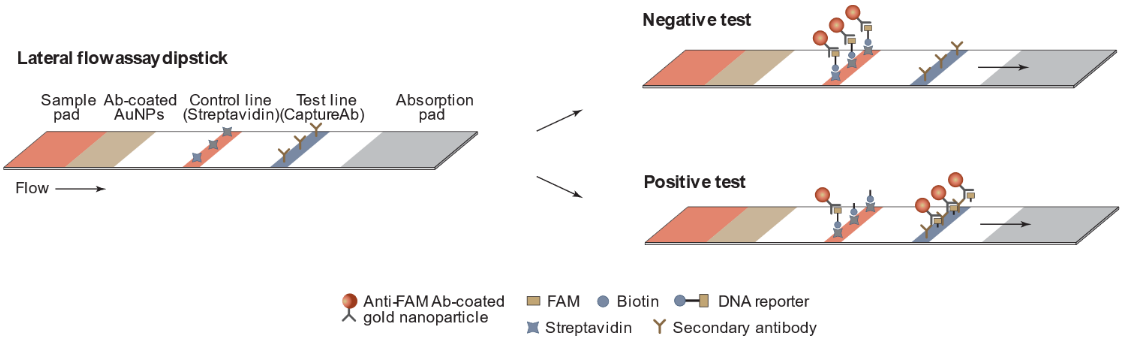


**Fig. S8. Principle of a lateral flow assay for visual detection of the SPEEDi-CRISPR product.** The control line captures biotin from both cleaved and uncleaved FB-ssDNA reporters. Activated RNA-coated beads cleave FB-ssDNA reporters, and FAM molecules from the cleaved reporters will be captured on the test line to produce signals.

**Table S1. Sequences appeared in this project.**

| Sequence Name | Sequence |
|---|---|
| ssDNA reporter | 56-FAM-TTATT-3IABkFQ |
| HPV 16 target | GTGGAACTTCACTTTTGTTAGCCTGTAATGTAGTAAAGTC |
| HPV 16 complementary | GACTTTACTACATTACAGGCTAACAAAAGTGAAGTTCCAC |
| HPV 18 target | CCTCACATTTAGTATCTTGCAATGTACTAAAGTCCATGGC |
| HPV 18 complementary | GCCATGGACTTTAGTACATTGCAAGATACTAAATGTGAGG |
| crRNA-1 | UAAUUUCUACUAAGUGUAGAUCGUCGCCGUCCAGCUCGACC |
| crRNA-HPV-16 | UAAUUUCUACUAAGUGUAGAUCUACAUUACAGGCUAACAAA |
| crRNA-HPV-18 | UAAUUUCUACUAAGUGUAGAUGUACAUUGCAAGAUACUAAA |
| PB-19 | CATTATTAAGTCCACTATTGTGGAAGCTGCAAAAGCTATT |
| HPV 18S | AACCTCTTCAGCACATGTTACTCTTAAGGAGTCTATGCAT |
| HPV 16S | TATTACCTTAACTAAGGCGTGATCTTAGGTTAGACGTTGT |

**Table S2. Recovery study of spiked serum sample.**

A

| Sample | Add (pM) | Found (pM) | Recovery (%) | RSD (%) |
|---|---|---|---|---|
| | 1.00 | 1.02 | 102 | 3.31 |
| HPV 18 | 0.100 | 0.106 | 106 | 4.44 |
| | 0.0100 | 0.00980 | 98.0 | 1.31 |
| | 1.00 | 0.938 | 93.8 | 5.04 |
| HPV 16 | 0.100 | 0.0997 | 99.7 | 1.73 |
| | 0.0100 | 0.0103 | 103 | 2.00 |

B

| Sample | Add (pM) | Found (pM) | Recovery (%) | RSD (%) |
|---|---|---|---|---|
| | 1.00 | 1.03 | 103 | 1.64 |
| HPV 18 | 0.100 | 0.102 | 102 | 1.69 |
| | 0.0100 | 0.0089 | 89 | 1.11 |
| | 1.00 | 0.99 | 99 | 3.90 |
| HPV 16 | 0.100 | 0.093 | 93 | 2.04 |
| | 0.0100 | 0.0101 | 101 | 4.34 |

**(A)** Recovery studies of SPEEDi-CRISPR detecting HPV 18 and HPV 16 in post-extraction spike serum samples. **(B)** Recovery studies of SPEEDi-CRISPR detecting HPV 18 and HPV 16 in heat-based lysis spiked serum samples (10%). Recovery rate = found/add x 100%.

**Table S3. Comparison of SPEEDi-CRISPR with other Cas12-based nucleic acid assays.**

| Name | Methods | CRISPR-Cas function | LOD | Time | Steps | Target | Amplification | Sample Processing | # of major components | Assay/Sensor Complexity |
|---|---|---|---|---|---|---|---|---|---|---|
| E-Si-CRISPR[1] | Electrochemistry | 1 function: Trans-cleavage | 3.5 fM | 100 min | 6 | dsDNA | No | Heat lysis | 1 enzymes, 2 probes, 2 other reagents, 1 sensor chip | High |
| SERS-CRISPR[2] | Electrochemistry | 1 function: Trans-cleavage | 1 fM | 50 min | 3 | RNA | No, RT needed | Column-based extraction | 2 enzymes, 4 probes | Moderate |
| (CRISPR)/Cas13a-powered biosensor[3] | Electrochemistry | 1 function: Trans-cleavage | 10 pM | 4 h | 3 | miRNA | No | Column-based extraction | 2 enzymes, 2 probes, 3 other reagents, 1 sensor chip | High |
| CRISPR–Chip[4] | Graphene field-effect transistor | 1 function: Targeting | 1.7 fM | 15 min | 2 | dsDNA | No | Column-based extraction | 1 enzyme, 2 probes, 1 sensor chip | High |
| ITP–CRISPR[5] | Fluorescence | 1 function: Trans-cleavage | 23 aM | 40 min | 3 | RNA | RT-LAMP | ITP extraction | 3 enzymes, 8 probes | High |
| dCas9-based solid-phase detection[6] | Fluorescence | 1 function: Targeting | 100 fM | 90 min | 4 | dsDNA | RPA and RCA | Chitosan-based extraction | 6 enzymes, 5 probes, 1 bead material | High |
| iPCCA[7] | Fluorescence | 1 function: Trans-cleavage | 1 fM | 140 min | 3 | RNA | No, primer extension needed | Column-based extraction | 2 enzymes, 4 probes | Moderate |
| SPEEDi-CRISPR | Fluorescence | 2 functions: Target enrichment, trans-cleavage | 2.3 fM | 80 min | 3 | dsDNA | No | Heat lysis | 1 enzyme, 2 probes, 1 bead material | low |

### Reference:


1. Suea-Ngam, A.; Howes, P. D.; deMello, A. J., An amplification-free ultra-sensitive electrochemical CRISPR/Cas biosensor for drug-resistant bacteria detection. *Chem. Sci.* **2021**, 12 (38), 12733-12743.

2. Liang, J.; Teng, P.; Xiao, W.; He, G.; Song, Q.; Zhang, Y.; Peng, B.; Li, G.; Hu, L.; Cao, D.; Tang, Y., Application of the amplification-free SERS-based CRISPR/Cas12a platform in the identification of SARS-CoV-2 from clinical samples. *J. Nanobiotechnology* **2021**, 19 (1), 273.

3. Bruch, R.; Baaske, J.; Chatelle, C.; Meirich, M.; Madlener, S.; Weber, W.; Dincer, C.; Urban, G. A., CRISPR/Cas13a-Powered Electrochemical Microfluidic Biosensor for Nucleic Acid Amplification-Free miRNA Diagnostics. *Adv. Mater.* **2019**, 31 (51), e1905311.

4. Hajian, R.; Balderston, S.; Tran, T.; deBoer, T.; Etienne, J.; Sandhu, M.; Wauford, N. A.; Chung, J. Y.; Nokes, J.; Athaiya, M.; Paredes, J.; Peytavi, R.; Goldsmith, B.; Murthy, N.; Conboy, I. M.; Aran, K., Detection of unamplified target genes via CRISPR-Cas9 immobilized on a graphene field-effect transistor. *Nat. Biomed. Eng.* **2019**, 3 (6), 427-437.

5. Ramachandran, A.; Huyke, D. A.; Sharma, E.; Sahoo, M. K.; Huang, C.; Banaei, N.; Pinsky, B. A.; Santiago, J. G., Electric field-driven microfluidics for rapid CRISPR-based diagnostics and its application to detection of SARS-CoV-2. *Proc. Natl. Acad. Sci. U S A* **2020**, 117 (47), 29518-29525.

6. Bengtson, M.; Bharadwaj, M.; Franch, O.; van der Torre, J.; Meerdink, V.; Schallig, H.; Dekker, C., CRISPR-dCas9 based DNA detection scheme for diagnostics in resource-limited settings. *Nanoscale* **2022**, 14 (5), 1885-1895.

7. Li, Y.; Mansour, H.; Watson, C. J. F.; Tang, Y.; MacNeil, A. J.; Li, F., Amplified detection of nucleic acids and proteins using an isothermal proximity CRISPR Cas12a assay. *Chem. Sci.* **2021**, *12* (6), 2133-2137.
